# Integrating More Walking into Public Transit Commuting: A Proof-of-Concept Study

**DOI:** 10.1101/2024.07.07.24310038

**Authors:** Yuval Hadas, Dan Emanuel Katz, Jonathan Rabinowitz

**Author notes:** Corresponding author: Jonathan Rabinowitz, PhD. Declarations of interest: none.

## Abstract

Regular physical activity is essential for maintaining and improving overall health, yet many individuals struggle to incorporate it into their daily lives. Previous studies, primarily based on simulations, have suggested that mode shifts in public transportation (PT) can increase walking, enabling PT commuters to meet some physical activity goals while traveling. However, current trip planning apps used by PT commuters do not prioritize walking as part of the journey. This proof-of-concept study investigates the potential for integrating more walking into commutes using PT, without compromising travel time, accessibility, or level of service. It represents the first phase of the “More Walking” population-based wellness initiative, which aims to enhance walking opportunities by modifying trip planning apps. We examined the impact of different walking thresholds on walking distance using a trip-planning algorithm using the home addresses of 2,149 commuters traveling to the same workplace in a Tel Aviv suburb. Our findings reveal that increasing walking thresholds either maintained or reduced total travel time, these results suggest that introducing a “more walking” option in trip planning could be a feasible way of incorporating physical activity and wellness as part of daily commutes.

## 1. BACKGROUND

Regular physical activity is essential for maintaining and improving overall health, yet many individuals struggle to incorporate it into their daily lives. Walking has important health benefits, a recent meta-analysis that included almost 50,000 persons found that taking more steps per day was associated with a progressively lower risk of all-cause mortality (1). For many people, walking is part of daily routine, it is frequently the last mile solution for commuters using public transportation (PT).

### 1.1 LITERATURE REVIEW

The health benefits of walking have been studied thoroughly. A short walk of 20 minutes each day can reduce the chance of early death by 25%. Walking helps prevent chronic diseases such as heart disease, type 2 diabetes, risk of stroke and some cancers (1-3). Walking is easy to include as part of everyday activities. More people walking also benefits the community, it enhances both actual and perceived safety of public spaces, reduces vehicle emissions and traffic congestion, stimulates local businesses and economic growth (4). Moreover, walking as a mode of transport reduces financial burden, leading to transport savings. Promoting walking gives people the option to move around without the need for a private vehicle. Accessibility to PT ensures that individuals have diverse and convenient mobility, contributing to a more inclusive and accessible urban environment (5).

Previous studies, based on simulations (6-15), and projections based on sampling of GPS data (16), have reported the potential for increased walking resulting from mode shifts in public transport. Previous studies did not consider the impact of variable walking distances on overall travel time. In the current study, we used the origins and the destination of actual commuters.We examined the effects of changing walking thresholds in trip planning on time spent walking as well as on overall travel time. The study is part of our quest to identify and promote simple, sustainable and impactful healthy lifestyle changes. Our approach is grounded in time-use epidemiology which explores effects of health-related time-use patterns in populations and optimizing distribution of time for population health (17).

Many PT users rely on trip planning apps to navigate their routes and manage fare payments. These apps do not prioritize walking as part of the journey. Popular trip planners, such as Google Maps, Moovit, Citymapper, Apple Maps, Bus Nearby (Hopon Mobility, Ltd) and OneBusAway, lack the options to adjust the maximum allowable walking distance or set a minimum desired walking threshold. Some apps, like efobus (efobus.com), allow adjusting maximum aerial distance as a very rough proxy for walking distance. Another group of app’s focus on helping walkers find the most inviting route (e.g., walkonomics.com 2010-2018) but do not include PT. However, including an upwardly adjustable walking distance parameter in the trip planning application could potentially provide the user with additional trip options, the potential of quicker and more reliable trips (less transfers), and improved wellbeing (18).

We conducted a proof-of-concept study with the aim of demonstrating that increasing walking thresholds among PT users is a practical and effective method for enhancing physical activity without extending travel time. This study marks the first phase of the ‘More Walking’ project, a population-based wellness initiative aimed at promoting walking by modifying trip planning apps to incorporate increased walking options. In the current phase reported in this paper, we present a proof-of-concept study in which we examined whether by increasing walking thresholds in a trip planning app commuters could include walking more in their commute and getting to their destination without adding travel time. Put simply, we examined whether one can leave home and get to work at the same time and include a walk.

## 2. METHODS

To test the concept that commuters could incorporate more walking without increasing travel time, we used Open Trip Planner (OTP) software (19). OTP is a tool that helps plan itineraries using public transportation, walking, biking, or other options. It uses data from publicly available maps and public transportation routes and schedules to plan efficient routes. Using OTP, we planned travel itineraries to work, using the addresses of the 2,149 employees sharing the same workplace, Bar-Ilan University, situated in a suburb of Tel Aviv. OTP allows adjusting limits on walking distance. We varied the amount of walking distance from 100 to 2500 meters. The research protocol was exempted from full review by the Human Subjects Institutional Review Board at Bar-Ilan University as the use of this data set which contained only a list of addresses without other identifying information, had been previously approved for use in another study.

The first step was to generate PT itineraries using OTP. The destination in every itinerary was Bar-Ilan University and the origins were the employees home addresses. By varying walking distance and number of stops, multiple itineraries for each employee’s commute were generated. Each itinerary included all trip legs, the start and finish stops, modes, ride/walk times, waiting times and distance. The travel itineraries generated by OTP were saved into a database for further analysis.

The second step was to eliminate itineraries that were not likely to be used. This was done in Python using the Hierarchical Non-Dominated Sorting (NHDS) Pareto Front Identification algorithm **(20)** which identifies optimal solutions based on multiple criteria. The algorithm first sorts all itineraries for each employee in ascending order by travel time and walking distance. Then it compares the first itinerary with all others, to make a rapid distinction between different itineraries to see whether a solution’s total travel time and walk time are greater than or equal to those of any other solution. If both criteria are met, the solution is considered inferior and removed from the selection pool. This filtering process helped to eliminate inferior trip options that passengers would be unlikely to choose. The remaining solutions, which ranged from 2 to 6 for each employee, were grouped by different walking thresholds.

Figure 1 shows the process of applying the algorithm and a route generated. The employees’ address (aka origin) and the university address (aka destination) were input to OTP. Using local maps and transportation data, OTP generated routes with the chosen relevant information, including the trip start time, travel time, distance, and other details of the itinerary. The extracted data was then used to generate a new file that stores the results.

**Figure 1.**
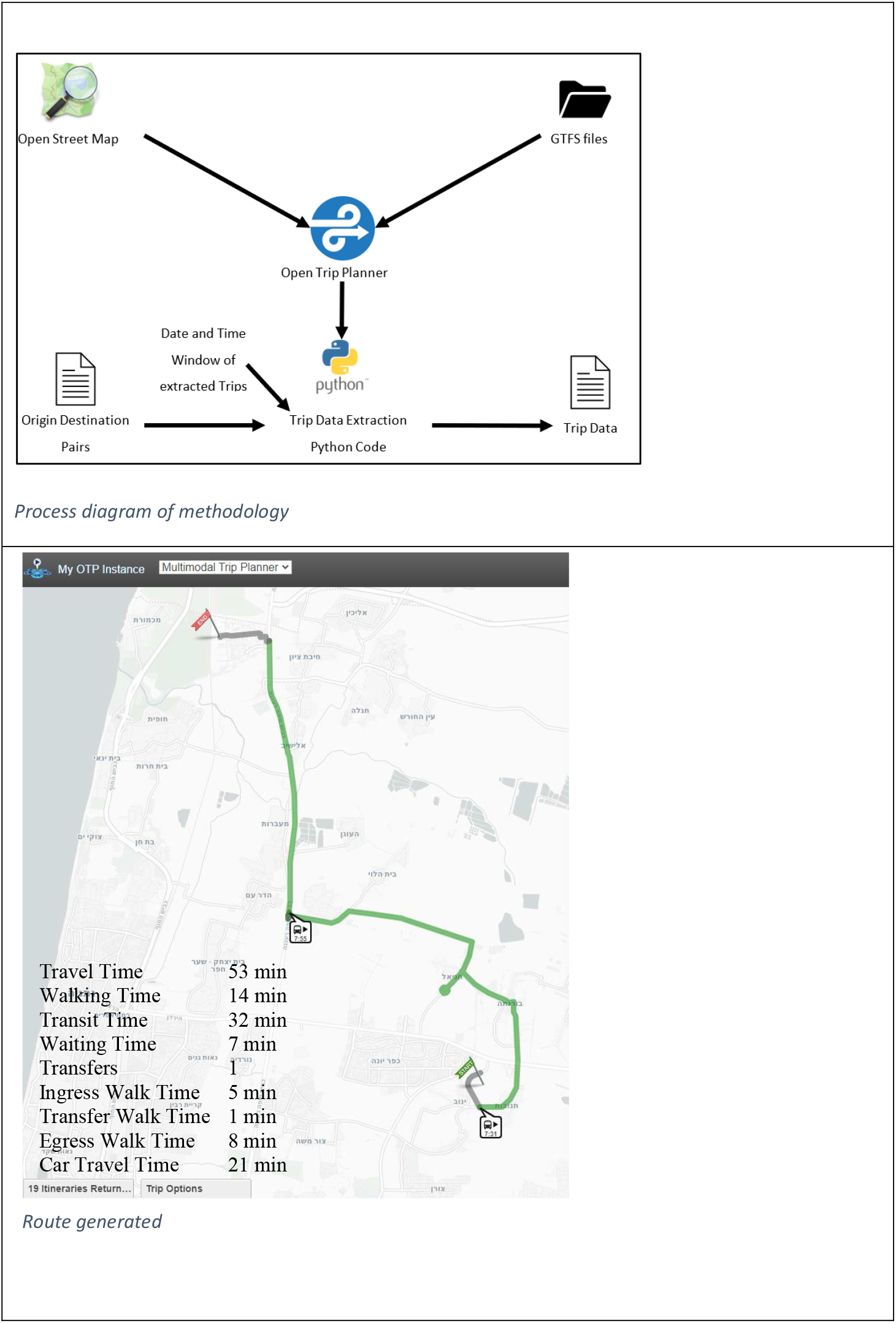
Process diagram and route generated.

Map in Figure 1 depicts a PT itinerary from OTP with key attributes. We used the following parameters presented in Figure 1 to analyze the impact of walking time and distance in the PT system on total travel time: (1) Time to reach the first stop from the origin (aka ingress time), (2) Transit time, (3) Transfer walk time, (4) Waiting time, and (5) Time to reach the destination from the last stop (aka egress time). The overall walking time includes walking time from the employees’ address to the boarding point, the transfer walking time (between transfer stations) and from the alighting point to destination. Transit time is the time spent traveling within the PT network. The number of transfers made during a trip indicates the complexity and convenience of interchanging between different PT modes or lines. Waiting time measures the time passengers spend waiting for the arrival of PT while transferring.

## 3.0 RESULTS

Figure 2 maps the location of all 2,149 employees. Each employee’s address is represented by a dot, which is colored based on the travel time by car to the campus. The yellow dot is the location of the campus. Data on potential itineraries travelling to work at Bar-Ilan University using public transportation were examined for various walking thresholds ranging from 100 to 2500 meters.

**Figure 2.**
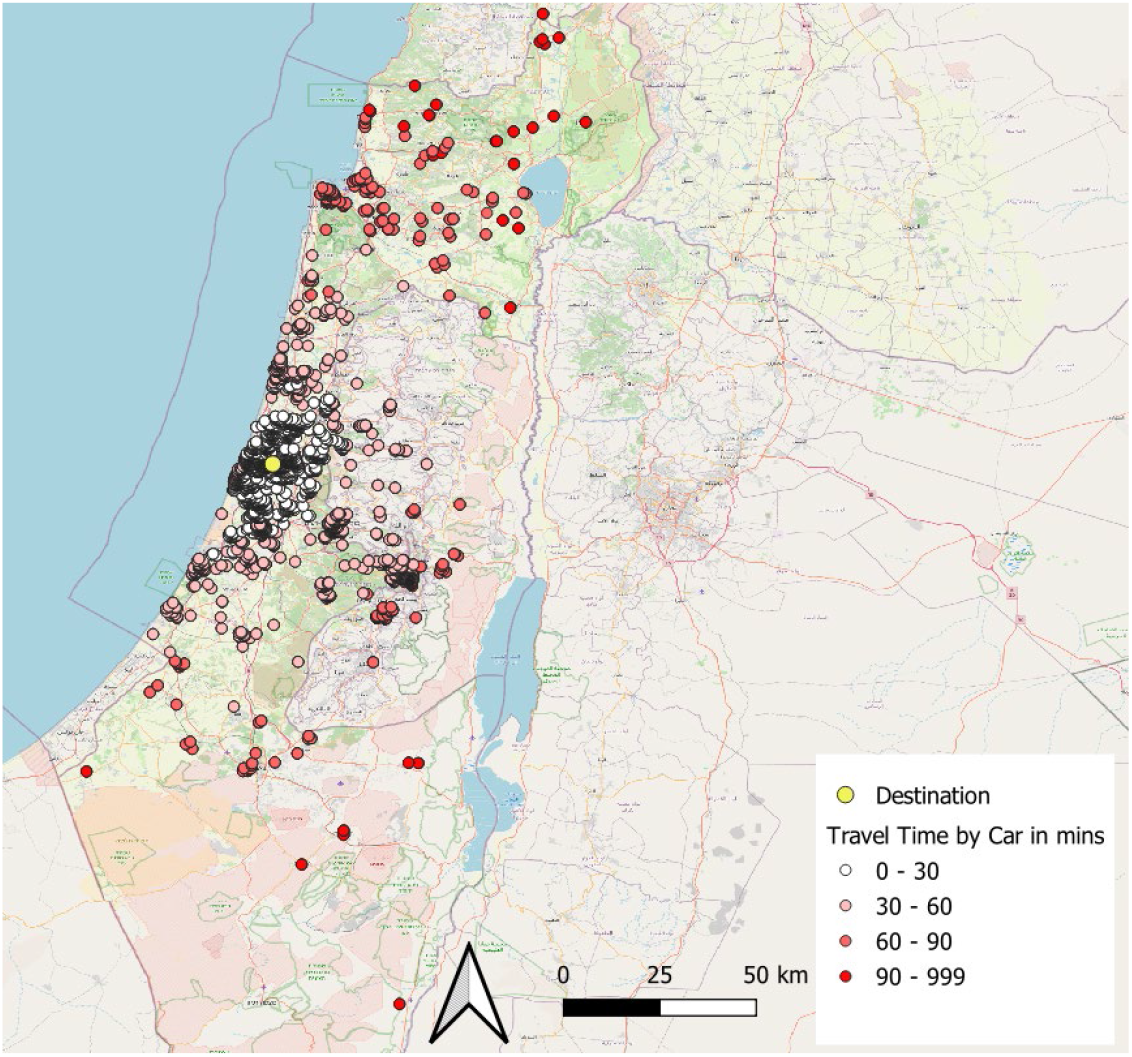
Origins (Bar-Ilan employees) and Destination (Bar-Ilan Campus) used in the case study.

Table 2 and Figure 3 present a descriptive summary by walking distance thresholds for all employees and for the subset of employees living within a 30-minute drive (Table 2 b, Figure 3, orange line). As seen in Table 2 (a & b) and Figures 3 (a, b & c), when more walking is introduced, there are more possible itineraries available, more origins are covered, meaning more employees can walk and use PT to get to work and less transfers are required. For example, only 33 employee addresses had routes requiring less than 100 meters walking whereas almost all of employees (2,135 out of 2,149) had routes requiring up to 2,500 meters of walking. For those living within 30 minutes by car travel (Table 2(b) and Figure 3 (d) orange line), transit time also declines as walking distance threshold increases. These decreases support the concept that the more passengers are willing to walk, the more direct and efficient the trip options become, reducing the need for transfer connections and waiting. Beyond the threshold of 1,000 meters of walking, the average number of transfers had only minor fluctuations. An additional advantage offered by the increased walking distance is that there are more stops that can be used upon arrival. There is one station at the entrance to the campus, 2 stations within 300 meters, 4 stations within 500 meters and 7 stations within 1000 meters.

**TABLE 2.**
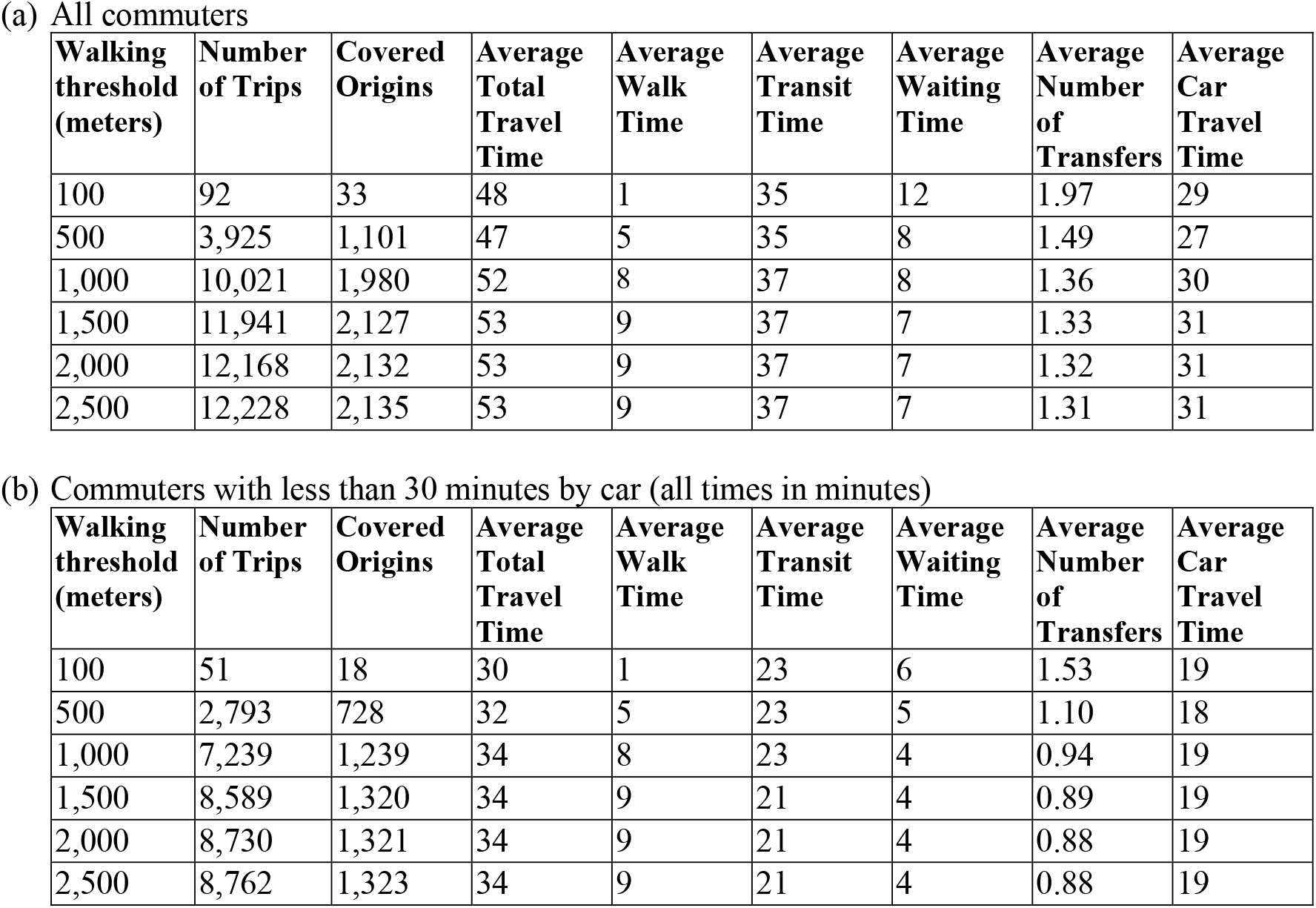
Descriptive summary by walking threshold (all times in minutes, rounded to nearest minute)

**Figure 3.**
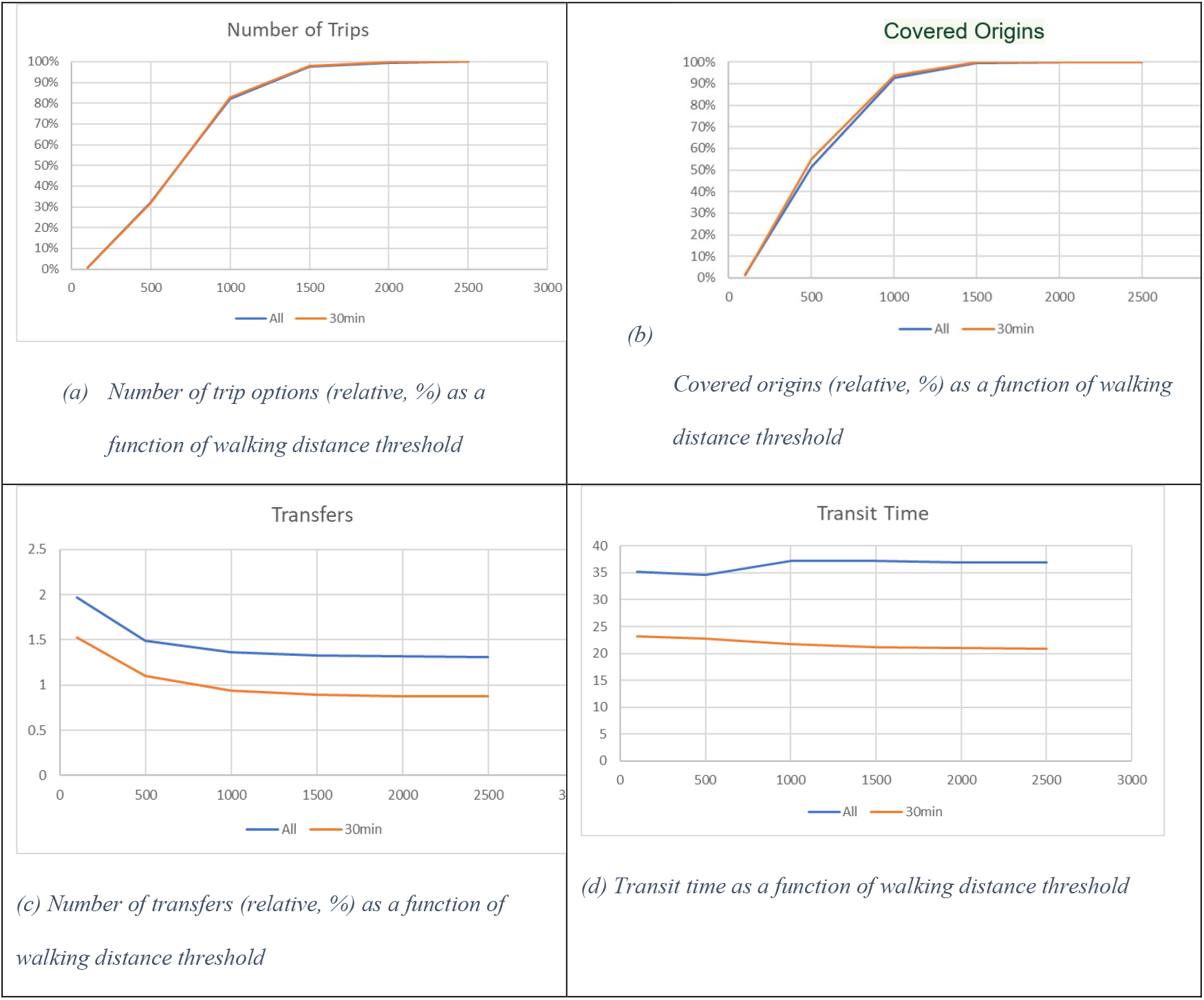
Effects of increasing walking thresholds (in meters) for all commuters (blue line) and those living within 30 minutes by car (orange line)

### 4.1. Discussion of results

As part of a population-based wellness intervention research initiative, we conducted a proof-of-concept study with the aim of examining the possibility that more walking could be included in commutes on public transportation. We did this by increasing walking thresholds in trip planning leveraging home address data on 2,149 employees working at the same location.We found that for most of the potential routes, as walking thresholds were increased, total travel time did not increase, and commuters were potentially able to benefit on average from a 9-minute walk going to work (walking threshold of 1500 meters). Considering that a short walk of 20 minutes each day can reduce the chance of early death by 25% (1-3) increasing walking thresholds as part of a daily commute has the potential to result in considerable health benefits. To further contextualize these findings we note that the United Nations has set in its Sustainability Development Guidelines a target of “Affordable and sustainable transport systems” (Target 11.2) which they define as convenient access to public transport within 500 meters to a stop or a station (https://sdgs.un.org/goals).

Previous research had used simulated data to demonstrate the concept that more physical activity could be included in commuting with public transportation (14, 21, 22). We have gone a step beyond demonstrating this by using real data to support the concept. These findings set the stage for our next study, which is a population-based intervention encouraging commuters to increase walking thresholds in their trip planning transportation apps.

### 4.2. Discussion of methods and limitations

Studies of transportation are dependent on infrastructures, services and locations. As such, each place differs, and the results based on this population may not be applicable to others. In this proof-of-concept study we were able to control destinations, as all people worked at the same location. Since each place differs all studies in this area are in some form or another case-studies. We believe that the findings from this case study are situationally grounded and generalizable meeting the duality principle for informative case research (23). Bar-Ilan University is in the greater Tel Aviv metropolitan area, with no special dedicated buses to the campus nor a train station. The results from this case are important as they provide additional evidence based on actual data of origin and a fixed destination that supports what other studies have found using simulations about the effects of modal switches in transit and walking. We have gone a step beyond, considering the effects of varying walking thresholds.

While the results are encouraging, we do not know the extent to which commuters will incorporate more walking; this will be explored in the next phase of the More Walking research program. Our results are limited as we were not able to take into consideration critical elements as noted in public policy transport guidelines as critical when attempting to integrate more walking into public transportation (24). These include elements such as safety, accessibility, how enjoyable walking routes are, public transport stops and stations, particularly for individuals with reduced mobility. Specifically, we did not take into account presence of infrastructure for weather protection, terrain mitigation, quality of street lighting, pedestrian-friendly crossings, wayfinding signage, obstacle-free pathways, sidewalk life and active frontages, need to carry cargo, possible difficulty walking long-distances and infrastructure to support individuals with disabilities. Finally, we did not take into consideration the risk of missing the service or missing a transfer or the effect of various walking paces. Future work in this area should strive to address the aforementioned and include measures of walkability (25-27).

## 5. Conclusions

A key potential benefit of incorporating more walking into public transportation (PT) commutes is the promotion of physical activity. By including walking segments in their journeys, commuters are more likely to engage in regular exercise, which can enhance their health and overall well-being. Integrating walking into daily commutes can play a significant role in fostering a healthier lifestyle.

The results of this proof-of-concept study suggest that increased walking as part of commute has the potential to result in shorter overall travel times and a larger variety of journey options for passengers. By having more choices for their routes, commuters can select paths that best suit their preferences and schedules. Additionally, this increase in journey options may enhance the overall reliability of the transportation system. When faced with unexpected delays or disruptions, passengers have alternative routes to choose from, this may minimize the negative impact on their daily commutes. Another benefit observed in the analysis is the reduction in the number of transfers and overall waiting time as walking distance increases. As passengers cover more distance on foot, the need for multiple transfers decreases. This translates to shorter waiting times at transfer points, contributing to a more time-efficient and convenient commuting experience. As a result, passengers can experience quicker and more efficient trips, making PT a more attractive option for daily commuting, increasing the potential that they will walk more daily.

In conclusion, this proof-of-concept study highlights the potential benefits of incorporating increased walking into public transportation (PT) commutes. Beyond the obvious health advantages, greater integration of walking within PT commutes could enhance reliability, expand journey options, and even improve overall time efficiency. Additionally, reducing transfers and waiting times can boost the effectiveness and convenience of the transportation system. By positioning walking as a key element of PT commutes, this approach shows promise in optimizing PT services while fostering healthier and more sustainable urban mobility. The next phase of this research will explore whether promoting increased walking can be effectively achieved by embedding walking thresholds into PT trip planning applications.

## Data Availability

All data produced in the present work are contained in the manuscript

## Declarations

### Ethics approval and consent to participate

Bar-Ilan University Institutional Review Board exempted the study from review and consent as this was as an analysis of an existing administrative data file, previously approved for use in a different study, and did not require collecting data from human subjects.

## Consent for publication

Not applicable.

## Availability of data and materials

Due to privacy concerns data can not be made available.

## Authors’ contributions

The authors confirm contribution to the paper as follows: study conception and design: YH, DK, JR; data collection: YH; analysis and interpretation of results: YH, DK; draft manuscript preparation: YH, DK, JR. All authors reviewed the results and approved the final version of the manuscript.

## Acknowledgments

This work was supported by a grant of the Israeli Smart Transportation

**Acknowledgments**

Not applicable.

